# The Impact of Glare Type and Intensity on Objective and Subjective Visual Performance

**DOI:** 10.1101/2025.04.18.25326064

**Authors:** Julian Hilmers, Maximilian Koschka, Eberhart Zrenner, Torsten Straßer

**Author notes:** **Corresponding author:** Dr. Torsten Straßer, Institute for Ophthalmic, Research Elfriede-Aulhorn-Str. 7, 72076 Tübingen, Germany. equal contribution (Eberhart Zrenner, Torsten Straßer). Disclosures: J.Hilmers, EyeServ GmbH (C); T.Straßer, EyeServ GmbH (C); M.Koschka, None; E.Zrenner, EyeServ GmbH (C), EyeServ GmbH (P).

## Abstract

**Purpose:** To compare the effects of spatially uniform glare and point light sources on visual acuity (VA) of individuals with normal vision under everyday contrast and illuminance conditions, and to investigate the related subjective perception of glare.

**Methods:** VA was assessed with spatially uniform glare (120 cm x 120 cm) lighting (VA-CAL test) and with four point light sources in either the paracentral (13.3°) or the near-peripheral visual field (23.5°). For all conditions, the illuminance and the optotype contrast (Landolt rings) were varied in four steps from 25 to 2200 lux and five steps from 20% to 80%, respectively. In addition to visual acuity testing, the subjective glare discomfort score was assessed for each illuminance using the Photoaversion Severity Questionnaire (PSQ).

**Results:** Twelve volunteers with normal vision (22 to 32 years, 6 women, 6 men) were enrolled in the study. Overall, regardless of the type of glare, increasing illuminance improved VA, while decreasing the contrast reduced VA. A statistically significant difference of 0.06 logMAR for the different types of glare was found only between uniform and point light glare in the near-periphery. The discomfort scores increased with illuminance, but did not differ statistically significantly between the different types of glare.

**Conclusions:** Both uniform and point light sources are a good way to measure daily visual performance in glare and show comparable results in influencing VA and subjective discomfort in everyday conditions.

**Translational Relevance:** Uniform light sources are just as suitable for testing visual acuity under glare as point light sources, although we recommend using uniform light for assessing daytime performance and punctual light for testing fitness to drive at night.

## INTRODUCTION

Visual acuity, as the most comprehensive and easily obtained parameter, plays an important role in the assessment of visual performance and is dependent on a number of factors, including contrast, ambient lighting, and glare. While these parameters are well defined for standardized acuity testing, in the real world these parameters are not constant, but are continually changing, as is visual acuity. Conventional visual acuity tests, such as the ETDRS chart [1], only measure visual acuity at a specific level of ambient light and at maximum contrast [2]. However, it has been shown that visual acuity improves with increasing luminance in individuals with normal vision [3,4]. In everyday life, our visual system must deal with a wide variety of light sources, ranging from small and bright point light glare such as oncoming car headlights or street lights at night [5, 6, 7], to large, uniform glare such as windows [8], reflective walls, street signs, sky on horizon, or sunlit snow patches in winter. Since the latter type of glare, point light glare, is of particular concern in traffic safety research, commercially available instruments such as mesoptometers or the nyctometers, can be used to test contrast vision in dim light with glare from point light sources [9]. In contrast, there are only a few studies on the effects of spatially uniform glare. Our group developed the Tuebingen VA-CAL test for measuring visual acuity under different levels of contrast and ambient luminance [10,11]. It uses a surface-emitting light source for a diffuse glare in the range of 25 to 2,200 lux as it can be found in everyday life, such as reflections of bright lights on walls, the overcast sky, road or information signs, either sunlit or self-illuminated. All of these lighting conditions share in common the necessity to allow the detection of different levels of contrast. We have recently found that many combinations of contrast and ambient luminance in daily living condition impact visual acuity in both healthy individuals and patients suffering from retinal disorders to various degrees [10,11]. To our knowledge, no study has directly compared the effects of point source and spatially uniform glare on visual acuity and subjective discomfort. This study examines how these different types of glare affect visual performance by assessing visual acuity at varying contrast levels and illuminance conditions representative of everyday life. Specifically, we compare point light sources, either paracentral or near-peripheral, with spatially uniform light sources. In addition, we investigate the effect of these light sources on subjective glare discomfort using the Photoaversion Severity Questionnaire.

## MATERIALS AND METHODS

### Experimental Setup

Visual acuity was determined at five contrast and six illuminance levels using a modified prototype (M&S Technologies, Niles, USA) implementing the VA-CAL-procedure described elsewhere [10]. For spatially uniform glare, four commercially available dimmable LED panels (each 60x60 cm, 60 W, 5500 – 6000 K; chromaticity: 0.313, 0.342; Prismica S.L., Valencia, Spain) were used, controlled using a computerized LED driver (IL-D53DALI2; Interlight GmbH, Arnsberg, Germany) to produce illuminances from 25 to 2,200 lux at the test distance of one meter. Optotypes (Landolt C rings) were presented on a high-resolution display (Microsoft Surface Pro 7; Microsoft Corporation, Redmond, Washington, United States), mounted in the center of the panels (Figure 1a). For point light a commercially available glare testing system (Glare Testing System, M&S Technologies, Niles, USA) was modified to include paracentral (13.3°) and the near-peripheral point light glare (23.5°). The CIE xy chromaticity of the LED spotlights is (0.454, 0.413), the illuminance ranges match those of LED panels (Figure 1b). To avoid reflections from the glare lights from walls or ceiling, black non-reflective folding screens were placed around the setups. For near-peripheral point light glare, the illuminance level was limited to 1400 lx due to technical limitations.

**Figure 1.**
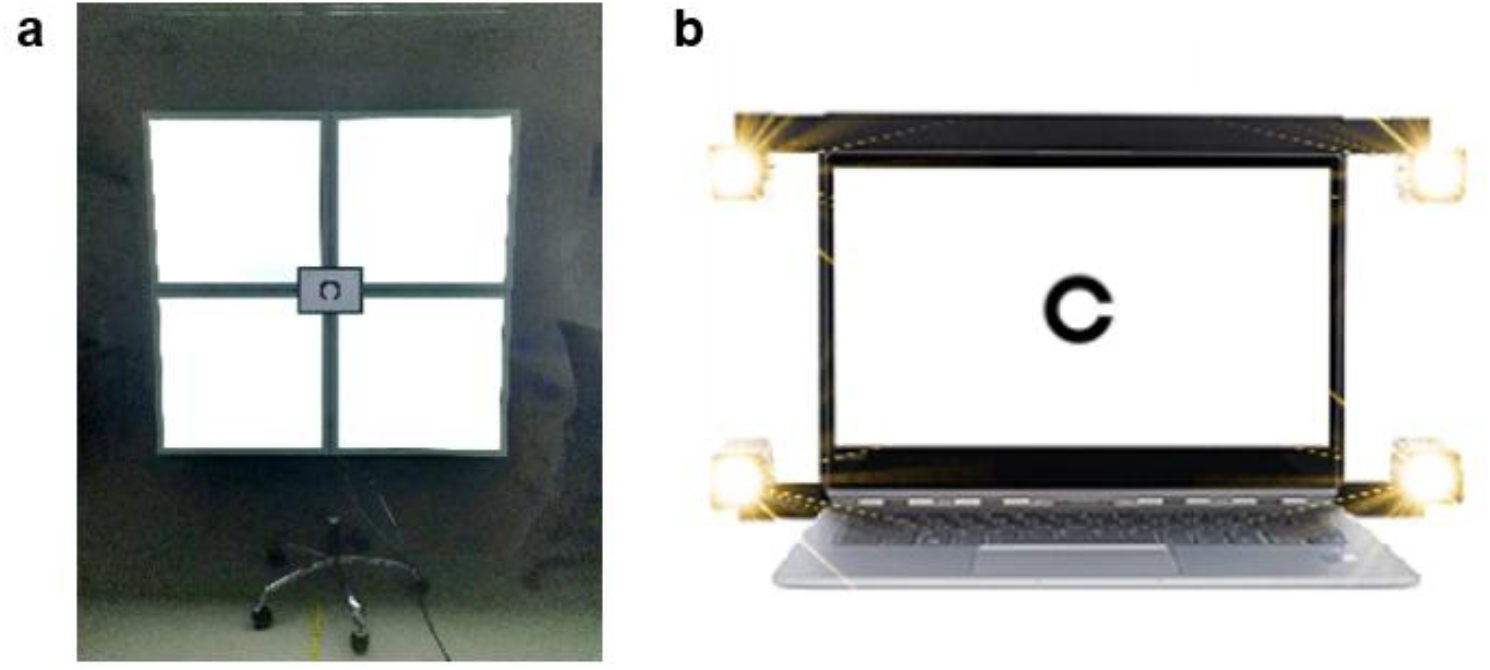
Experimental setup. a) Photograph of the VA-CAL prototype featuring four LED panels and a high-resolution screen for presenting optotypes; b) Sketch of the modified CTS VA testing system with four point light LEDs for paracentral glare.

### Study Participants

Twelve volunteers (6 female, 6 male) aged 22 to 32 years (mean ± SEM: 28.1 ± 2.7 years) were recruited from the student body of the University of Tübingen and the staff of the University Eye Hospital Tübingen and enrolled in the study after confirmation of the inclusion and exclusion criteria. Best-corrected visual acuity (BCVA) was assessed using a standardized ETDRS-chart (4 m, 100 cd/m^2^, 98% contrast) in a preliminary examination to ensure the inclusion criterion of a monocular visual acuity of at least 0.1 logMAR. Participants with ocular diseases were excluded. The study adhered to the tenets of the Declaration of Helsinki and was approved by the Ethics Committee of the Faculty of Medicine, University of Tübingen (431/2019BO2). All participants received detailed information about the study and provided written informed consent.

### Experimental Procedure

Participants were best corrected with an addition of +0.75 diopter to correct for near distance testing and were seated in a darkened room at a distance of one meter in front of the stimulation devices in a combined chin and head rest. After an initial 5-minute dark adaptation period during which instructions were given, five levels of illumination were presented in ascending order of intensity. A one-minute adaptation period was added before each illumination level, during which the participants were asked to fixate a red fixation cross (2.9°) in the center of the screen and to rate their subjective discomfort with the current glare condition between 0 and 10 (0 = no discomfort/pain; 10 = unbearable discomfort/pain) using the Photoaversion Severity Questionnaire, an extended version of the de Boer rating scale for discomfort glare [12]. At each illumination level, Landolt C rings of varying size were presented monocularly, and their size was controlled by an adaptive staircase procedure, while their orientation was randomly chosen from eight possible directions. Depending on the illumination level, the procedure was repeated for either two or three contrast levels (Weber 80%, 65%, 50%, 35% or 20%; Fig. 2), respectively, resulting in a total 13 visual acuity values. Participants had to indicate the direction of the opening of the Landolt C ring using a wireless keypad within a maximum of seven seconds. Failure to respond within this time or providing an incorrect answer was counted as a wrong response (8-alternative forced-choice). If the direction could not be identified, the participants were instructed to guess the direction. An auditory feedback was provided. The procedure ends, when the maximum attainable visual acuity is determined [13].

**Figure 2.**
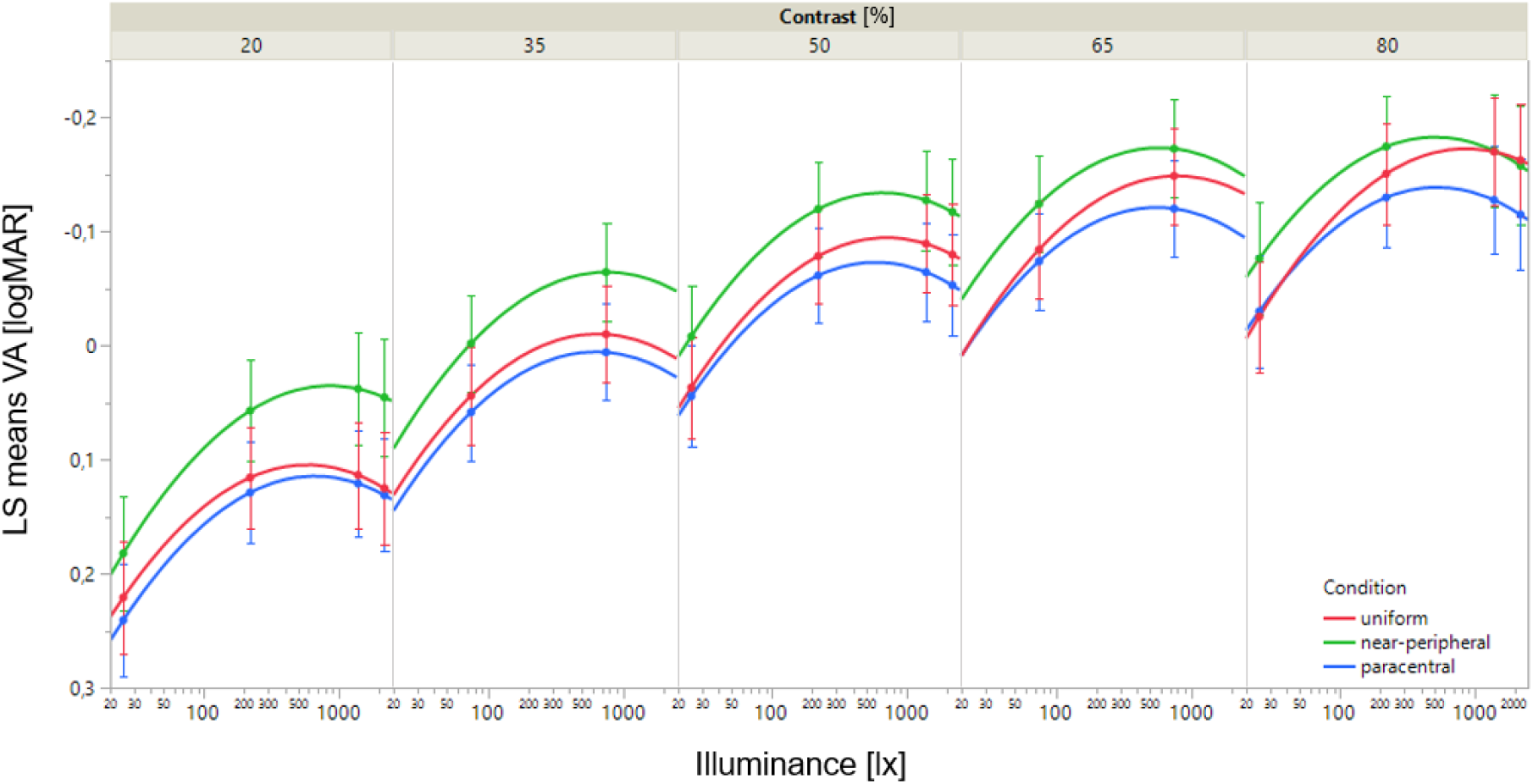
Visual acuity as a function of different types of glare, illuminance, and contrast as predicted from the linear mixed-effects model. With LS (Least Square) means VA [logMAR] on y-axis and illuminance on x-axis. The VA curves of the individual contrasts (20-80%) are shown separately and indicated by columns in ascending order from left to right. The different colors of the curves represent the different conditions (red = uniform glare, green = near-peripheral point light glare, blue = paracentral point light glare. The error bars indicate the 95% confidence interval at the tested combinations of illuminance and contrast.

Participants were randomly assigned to begin the experiment with either spatially uniform glare or point light glare. For point light glare, the paracentral glare condition was performed first, followed by the near-peripheral glare condition.

### Statistical analysis

To investigate the effects of the different glare types on VA under different levels of contrast and ambient luminance, a full-factorial linear mixed-effects model was employed, with the fixed factors being condition (light sources/levels: paracentral, near-peripheral, uniform glare), the continuous contrast level, and the continuous logarithmized illuminance, both modeled as quadratic terms. The participant was treated as a random effect. The model was fitted using restricted maximum likelihood (REML). The variance inflation factors (VIF) of the predictors were calculated and assured to fall well below the common threshold value, indicating no collinearity between them [14]. The models’ residuals were confirmed visually to follow a normal distribution and the homogeneity of the variances was ensured using the Brown– Forsythe test and reported in case of violations [15,16]. Post hoc comparisons of the least-squares means using two-tailed t-tests were conducted in case of statistically significant effects. One participant was unable to take part in the peripheral measurement for personal reasons, meaning that only 11 people were considered here.

The participants’ discomfort score regarding the Photoaversion Severity Question were analyzed using a full-factorial generalized mixed model with ordered beta regression [17] (estimated using maximum likelihood), with the fixed effects glare condition (light sources/levels: paracentral, near-peripheral, uniform glare) and illuminance (continuous). To account for repeated measurements and unequal group sizes the participant was included as random effect.

Statistical analyses were performed using JMP Pro 17 (SAS Institute, Cary, United States) or the R Statistical language (version 4.2.2; [18]), using the packages marginaleffects (version 0.14.0; [19]) and glmmTMB (version 1.1.7; [20]). If not otherwise stated, an alpha level of 0.05 was used for all statistical analyses.

## RESULTS

The mean participants’ BCVA determined with the standard ETDRS-chart (4 m; luminance = 100 cd/m^2^; contrast = 98%) was between -0.1 and -0.3 logMAR (mean ± SEM: -0.18 ± 0.10 logMAR).

Figure 2 shows how visual acuity depends on the different glare types (uniform, near-peripheral, paracentral) at different illuminances and contrasts. The pattern of least-square means VA as a function of illuminance and contrast was similar for all glare types (Figure 2). Increasing the illuminance up to approximately 750 lx improved the VA on average, while decreasing the contrast deteriorated mean VA regardless of illuminance. The linear mixed-effects model revealed statistically significant effects of illuminance, contrast (both p < 0.0001), and type of glare (p = 0.0482) on VA as shown in Table 1. A post hoc paired t-test revealed a statistically significant mean difference of 0.06 logMAR between uniform glare and near-peripheral point light glare (Table 2), but not between paracentral and near-peripheral point light as well as uniform light and paracentral point light.

**Table 1.** Results of the linear mixed-effects model (n = 490, R2adj. = 0.8133) with the dependent variable visual acuity and the random effect participant.

| Effect | F-statistic | p-value |
| --- | --- | --- |
| Log(Illuminance) | $F(1, 465) = 100.5065$ | $<.0001^{***}$ |
| Contrast | $F(1, 465) = 101.2676$ | $<.0001^{***}$ |
| Glare type | $F(2, 465) = 3.0513$ | $.0482^*$ |
| Log(Illuminance)*Contrast | $F(1, 465) = 0.8239$ | $.3645$ |
| Log(Illuminance)* Glare type | $F(2, 465) = 0.7464$ | $.6047$ |
| Contrast*Glare type | $F(2, 465) = 1.4713$ | $.3894$ |
| Log(Illuminance)*Contrast*Glare type | $F(2, 465) = 2.2823$ | $.1687$ |
| Log(Illuminance)*Log(Illuminance) | $F(1, 465) = 72.3179$ | $<.0001^{***}$ |
| Contrast*Contrast | $F(1, 465) = 136.2783$ | $<.0001^{***}$ |

Alpha level = 0.05; asterisks indicate the level of significance: \* $p < .05$ , \*\* $p < .01$ , \*\*\* $p < .001$

**Table 2.** Results of post hoc comparisons using paired t-tests of the least-squares means obtained from the linear mixed-effects model. Each glare type was compared with another and the mean VA difference between them was calculated.

| Comparison | Difference [95% CI] (logMAR) | t-statistic | p-value |
| --- | --- | --- | --- |
| uniform – near-peripheral | 0.06 [0.01, 0.11] | $t(465.0595) = 2.3485$ | $.0193^*$ |
| paracentral – near-peripheral | 0.05 [0.00, 0.10] | $t(465.0595) = 1.9492$ | $.0519$ |
| uniform - paracentral | 0.01[-0.04, 0.06] | $t(465.0171) = 0.4255$ | $.6707$ |

Alpha level = 0.05; asterisks indicate the level of significance: \* $p < .05$ , \*\* $p < .01$ , \*\*\* $p < .001$

Figure 3 shows the glare discomfort ratings obtained from the Photoaversion Severity Questionnaire (y-axis) for different types of glare (colored lines) and illuminance levels (x-axis). As expected, the generalized linear mixed model revealed a statistically significant effect of illuminance on discomfort scores (Table 3), with higher illuminance levels leading to greater discomfort regardless of the type of glare. For all illuminances, the uniform light source was rated as the most comfortable, followed by the near-peripheral and paracentral point sources. However, there was no statistically significant effect between the different types of glare in terms of discomfort.

**Figure 3.**
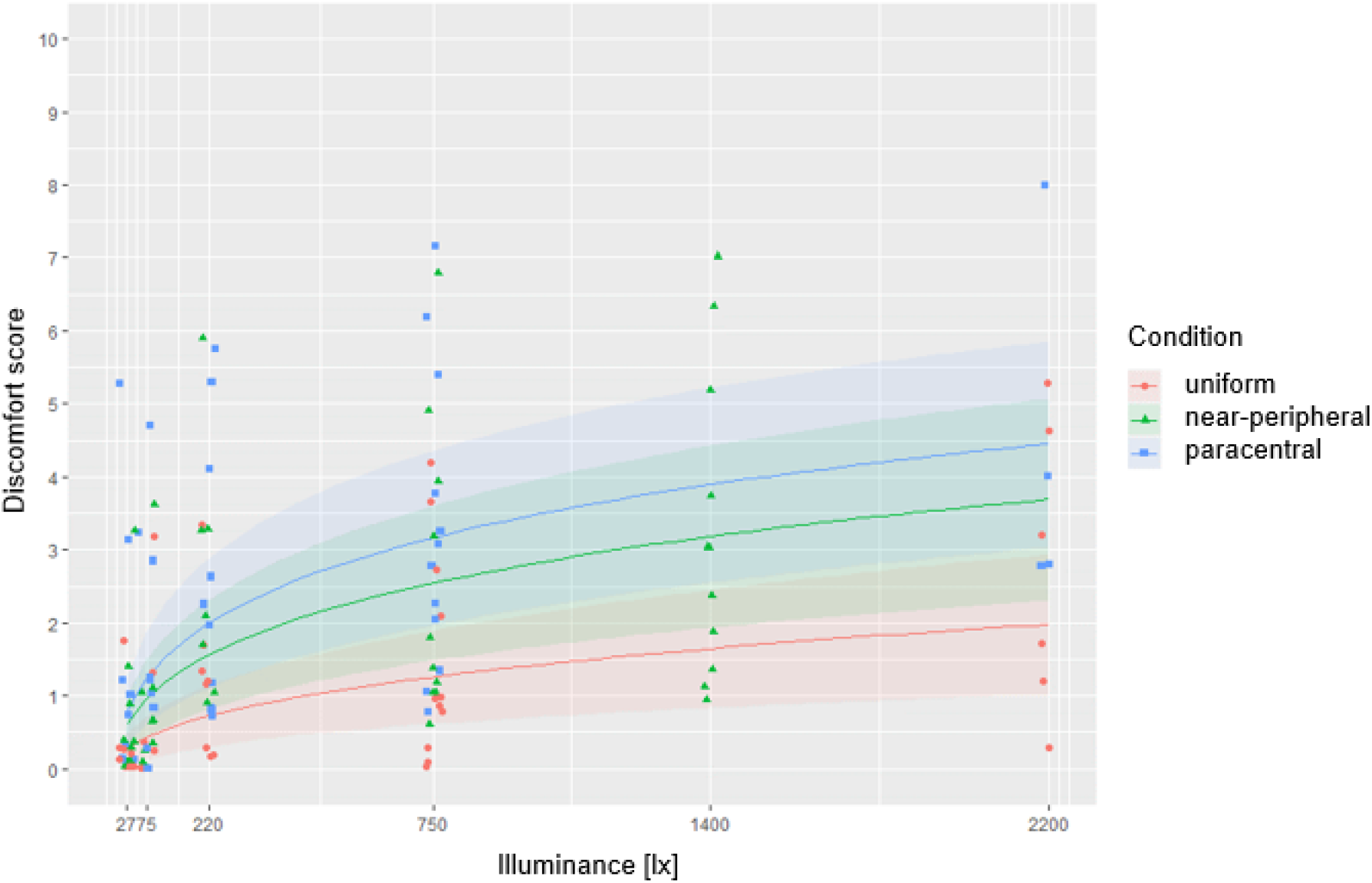
Discomfort scores of the Photoaversion Severity Questionnaire as a function of illuminance and glare type (symbols) and prediction of the ordered beta regression (lines). Colors represent the different types of glare, shaded bands 95% confidence intervals of the prediction.

**Table 3.** Results of the generalized linear mixed model with ordered beta regression (n = 175, dispersion parameter = 21.4). The model’s intercept corresponds to uniform glare at an illuminance of 0 lx.

| Effect | Estimate (SE) | z-Value | p-value |
| --- | --- | --- | --- |
| (Intercept) | -5.253 (0.670) | -7.836 | <.001*** |
| paracentral | 0.874 (0.720) | 1.214 | .225 |
| near-peripheral | 1.123 (0.669) | 1.680 | .093 |
| log(Illuminance) | 0.501 (0.090) | 5.560 | <.001*** |
| paracentral*log(Illuminance) | -0.002 (0.110) | -0.015 | .988 |
| near-peripheral*log(Illuminance) | 0.007 (0.007) | 0.068 | .946 |

Alpha level = 0.05; asterisks indicate the level of significance: \* $p < .05$ , \*\* $p < .01$ , \*\*\* $p < .001$

## DISCUSSION

In a previous study, we demonstrated that visual acuity is not constant and represented by a single value [4], as suggested by the well-established clinical standard [2]. Instead, visual acuity varies continuously with contrast and illuminance [4]. Conventional visual acuity testing, in which visual acuity is assessed at maximum contrast at a given illuminance [1,2], is therefore unable to describe visual acuity in everyday situations with constantly changing contrast and illuminance conditions and is essentially limited to an artificial condition. Visual acuity decreases especially under glare and low contrast conditions [3,4], which may lead to overestimation of visual acuity, especially in patients who are sensitive to glare [11]. In this study we investigated the effect of different glare types and varying contrasts on visual acuity (disability glare), and on subjective glare perception (discomfort glare). Contrary to our intuitive hypothesis that point light glare would lead to both greater discomfort and increased disability glare, our results did not generally confirm this. While no statistically significant difference was found between the different types of glare on the Photoaversion Severity Questionnaire discomfort score, a small but statistically significant reduction in visual acuity was found for near-peripheral point source glare (23.5°) compared to uniform glare, but not between near-peripheral and paracentral (13.3°) or paracentral and uniform glare. Interestingly, the increase in disability glare seems to have no effect on discomfort glare in our young population of volunteers, although Lin and colleagues describe a correlation between glare angle and subjective glare perception for angles ≥ 10° [21].

One would expect that a uniform diffuse light source would illuminate the retina more evenly than point sources, which stimulate only small areas of the retina, resulting in increased discomfort and disability glare. However, the results of this study contradict this hypothesis in young healthy adults, at least for the particular conditions tested. Further factors influencing glare in addition to the light intensity and shape (uniform vs. point source) of the light sources are the size and number of light sources, the position in the field of vision and the adaptation state of the retina [22].

Sivak et al. [23] showed that the area of the light source affects glare discomfort: smaller light sources produce the same brightness over a smaller area, so the brightness is locally more intense. However, point sources of light are not imaged as “dots” on the photoreceptor mosaic; rather, the light is distributed across the retina by diffraction and imperfections in the eye’s optics and scattering [24]. Future research needs to focus on possible differences in the effects of uniform and point glare on disability glare and, in particular, discomfort glare in patients with cataracts, as these are known to show significant disability glare and reduced contrast sensitivity [25], which affects their fitness to drive [26], among other problems.

The area and shape of the light source also affects the retina’s state of adaptation [24, 27]. A large uniform light source is likely to provide better light adaptation than comparably small point light sources, especially considering that intrinsically photosensitive retinal ganglion cells (ipRGCs) are non-image-forming encoders of ambient light [28]. However, multiple point light sources in the field of view may counteract this effect, possibly explaining the absence of differences between glare conditions in our study. In nighttime road traffic, multiple point light sources contribute to visual interference, particularly from oncoming vehicles. Conventional glare tests typically employ only a single point light source [9], possibly resulting in increased discomfort and disability glare. It should also be noted that previous studies investigating discomfort glare were conducted under scotopic or mesopic conditions, with background luminance limited to 3 cd/m^2^ [29, 30], or under low-luminance photopic conditions, with ambient luminance limited to < 100 cd/m^2^ [27], and therefore their results are not directly comparable to ours, as studies at high luminance levels are missing.

In addition to the number of point sources, their position in the visual field also plays an important role. Interestingly, as mentioned above, Lin et al. found that a glare source at 10° from the line of sight caused slightly more discomfort than a source at 20° [21], whereas in our study we found no difference in either discomfort glare or disability glare between near-peripheral (23.5°) and paracentral (13.3°) point light sources. Jones et al. describe a reduction in contrast sensitivity under scotopic glare conditions (0.32 cd/m^2^) for a point light source of 0.25° diameter at 3° eccentricity [31], but did not compare different eccentricities. In a study comparing high intensity discharge (HID) and halogen head lamps, Bullough and colleagues found statistically significant effects of eccentricity (5° vs. 10°) on both discomfort and disability glare (measured as contrast sensitivity) independent of lamp type, with worse values for the smaller eccentricity [32] Further research should investigate the effect of glare light source eccentricity on both glare discomfort and disability, especially in the central visual field. To our knowledge, no studies have addressed this aspect.

Bullough et al. also report a statistically significant effect of lamp type on glare discomfort, namely stronger discomfort ratings with HID lamps, which is white-blue, compared to halogen and blue-filtered halogen lamps, which are rather yellowish, independent of the illuminances [32]. They attributed the greater discomfort to the higher energy in the short-wavelength range of the visible spectrum, where blue cones are most sensitive. Interestingly, this seems to have no effect on glare disability [32]. These findings were confirmed by Flannagan et al. who found the lowest discomfort ratings for yellowish light sources with a peak wavelength of 577 nm [29, 33].

However, the results of Bullough et al. and Flannagan et al. also point to a limitation of our study, which used different color temperatures: the uniform glare is produced by white-blueish LEDs behind a diffusing screen, while the point light sources produce a warm, yellowish light. This difference in color temperature may also partially explain the results of our study and should be investigated further.

Another limitation of our study is the young, ocularly healthy study population. Older individuals may exhibit a greater effect of point light sources on visual acuity because age-related factors such as dry eye disease and lens opacities (e.g., cataracts) can increase light scattering [34-36]. Similarly, certain clinical conditions, such as achromatopsia [37, 38] and congenital stationary night blindness [39], are associated with cone dysfunction and pronounced photophobia, which may exacerbate glare sensitivity. While previous research has demonstrated impairment in achromats under uniform lighting conditions [11], their response to point light sources remains unstudied. In addition, individual differences in light perception were observed in our study. Increasing the sample size may help to account for such variability. Therefore, further research is needed, especially with different groups of participants.

In summary, both glare conditions are necessary to fully evaluate visual performance in everyday life. While VA-CAL with uniform glare primarily simulates daylight conditions, point light sources at different viewing angles replicate nighttime glare scenarios such as oncoming headlights. Both types of glare result in reduced visual acuity and increased discomfort beyond a certain illuminance threshold. Because photosensitive individuals are often unlicensed drivers and experience the greatest impairment during daylight hours, we recommend using a uniform light source to assess their daily visual function. In contrast, nighttime driving ability should continue to be assessed using point light sources. With the advent of coherent laser light sources in automotive headlights [40] and the widespread use of pulse width modulation (PWM) for brightness control in LED headlights [41], glare characteristics are evolving [42]. Coherent laser light sources can produce highly collimated beams, potentially increasing the intensity and sharpness of glare effects, while PWM modulation introduces flicker, which may further contribute to visual discomfort, particularly in photosensitive individuals [43]. Future studies should take these technological advances into account when evaluating glare effects. In addition, point light sources should be placed more centrally than the 23.5° and 13.3° eccentricities used in this study to ensure an adequate glare effect under test conditions.

## Data Availability

All data produced in the present study are available upon reasonable request to the authors.

## ACKNOWLEDGMENTS

The authors would like to express thanks to the Tistou and Charlotte Kerstan Foundation 2000 for the financial support. Further, we acknowledge the support of the Open Access Publication Fund of the University of Tübingen. We would also like to thank all the volunteers who took part in this study.

## REFERENCES

[1] Shamir, R. R., Friedman, Y., Joskowicz, L., Mimouni, M., & Blumenthal, E. Z. (2016). Comparison of Snellen and Early Treatment Diabetic Retinopathy Study charts using a computer simulation. International journal of ophthalmology, 9(1), 119–123.

[2] Wesemann, W., Heinrich, S. P., Jägle, H., Schiefer, U., & Bach, M. (2020). New DIN and ISO norms for determination of visual acuity. Der Ophthalmologe, 117, 19–26.

[3] Hauser, B., Ochsner, H., & Zrenner, E. (1992). Der "Blendvisus”-Teil 1: Physiologische Grundlagen der Visusänderung bei steigender Testfeldleuchtdichte. Klinische Monatsblätter für Augenheilkunde, 200(02), 105–109.

[4] Hilmers, J., Straßer, T., Bach, M., Stingl, K., & Zrenner, E. (2022). Quantification of the dynamic visual acuity space at real-world luminances and contrasts: the VA-CAL test. Translational Vision Science & Technology, 11(4), 12–12.

[5] Hu, J., Guo, Y., Wang, R., Ma, S., & Yu, A. (2022). Study on the influence of opposing glare from vehicle high-beam headlights based on drivers’ visual requirements. International journal of environmental research and public health, 19(5), 2766.

[6] Gil, P. A., De Santos-Berbel, C., & Castro, M. (2021). Driver glare exposure with different vehicle frontlighting systems. Journal of safety research, 76, 228–237.

[7] Wood, J. M., Isoardi, G., Black, A., & Cowling, I. (2018). Night-time driving visibility associated with LED streetlight dimming. Accident Analysis & Prevention, 121, 295–300.

[8] Osterhaus, W. K. (2005). Discomfort glare assessment and prevention for daylight applications in office environments. Solar Energy, 79(2), 140–158.

[9] Aulhorn, E., & Harms, H. (1965). The mesoptometer, a device for the examination of twilight vision and susceptibility to blinding. Bericht über die Zusammenkunft. Deutsche Ophthalmologische Gesellschaft, 66, 425–426.

[10] Hilmers, J., Straßer, T., Bach, M., Stingl, K., & Zrenner, E. (2022). Quantification of the dynamic visual acuity space at real-world luminances and contrasts: the VA-CAL test. Translational Vision Science & Technology, 11(4), 12–12.

[11] Hilmers, J., Bach, M., Stingl, K., Zrenner, E., & Straßer, T. (2023). The VA-CAL test quantifies improvement of visual acuity in achromatopsia by means of short-wave cutoff filter glasses in daily living conditions. Translational Vision Science & Technology, 12(6), 20–20.

[12] Gellatly, A. W., & Weintraub, D. J. (1990). User reconfigurations of the de Boer rating scale for discomfort glare. University of Michigan, Ann Arbor, Transportation Research Institute.

[13] Beck, R. W., Moke, P. S., Turpin, A. H., Ferris III, F. L., SanGiovanni, J. P., Johnson, C. A., … & Kraker, R. T. (2003). A computerized method of visual acuity testing: adaptation of the early treatment of diabetic retinopathy study testing protocol. American journal of ophthalmology, 135(2), 194–205.

[14] Hair, J.F.J.; Anderson, R.E.; Tatham, R.L.; Black, W.C. Multivariate Data Analysis; 3^rd^ ed.; Macmillan: New York, 1995;

[15] Santos Nobre, J., & da Motta Singer, J. (2007). Residual analysis for linear mixed models. Biometrical Journal: Journal of Mathematical Methods in Biosciences, 49(6), 863–875.

[16] Crosby, J. M., Twohig, M. P., Phelps, B. I., Fornoff, A., Boie, I., Mazur-Mosiewicz, A., … & Allen, R. L. (2011). Homoscedasticity. Encyclopedia of Child Behavior and Development, 752–752.

[17] Kubinec, R. (2023). Ordered beta regression: a parsimonious, well-fitting model for continuous data with lower and upper bounds. Political Analysis, 31(4), 519–536.

[18] R Core Team, R. (2013). R: A language and environment for statistical computing.

[19] Arel-Bundock, V., Greifer, N., & Heiss, A. (2024). How to interpret statistical models using marginaleffects for R and Python. Journal of Statistical Software, 111, 1–32.

[20] Brooks, M. E., Kristensen, K., Van Benthem, K. J., Magnusson, A., Berg, C. W., Nielsen, A., … & Bolker, B. M. (2017). glmmTMB balances speed and flexibility among packages for zero-inflated generalized linear mixed modeling. The R journal, 9(2), 378–400.

[21] Lin, Y., Liu, Y., Sun, Y., Zhu, X., Lai, J., & Heynderickx, I. (2014). Model predicting discomfort glare caused by LED road lights. Optics express, 22(15), 18056–18071.

[22] Wang, S., & Zhao, J. (2022). New prospectives on light adaptation of visual system research with the emerging knowledge on non-image-forming effect. Frontiers in Built Environment, 8, 1019460.

[23] Sivak, M., Simmons, C. J., & Flannagan, M. (1990). Effect of headlamp area on discomfort glare. Lighting Research & Technology, 22(1), 49–52.

[24] Shevell, S. K., & Burroughs, T. J. (1988). Light spread and scatter from some common adapting stimuli: Computations based on the point-source light profile. Vision Research, 28(5), 605–609.

[25] Rubin, G. S., Adamsons, I. A., & Stark, W. J. (1993). Comparison of acuity, contrast sensitivity, and disability glare before and after cataract surgery. Archives of Ophthalmology, 111(1), 56–61.

[26] Bal, T., Coeckelbergh, T., Van Looveren, J., Rozema, J. J., & Tassignon, M. J. (2011). Influence of cataract morphology on straylight and contrast sensitivity and its relevance to fitness to drive. Ophthalmologica, 225(2), 105–111.

[27] Bargary, G., Jia, Y., & Barbur, J. L. (2015). Mechanisms for discomfort glare in central vision. Investigative ophthalmology & visual science, 56(1), 464–471.

[28] Do, M. T. H., & Yau, K. W. (2010). Intrinsically photosensitive retinal ganglion cells. Physiological reviews.

[29] Flannagan, M. J., Sivak, M., & Gellatly, A. W. (1991). Joint effects of wavelength and ambient luminance on discomfort glare from monochromatic and bichromatic sources. University of Michigan, Ann Arbor, Transportation Research Institute.

[30] Weintraub, D. J., Gellatly, A. W., Sivak, M., & Flannagan, M. J. (1991). Methods for evaluating discomfort glare. University of Michigan, Ann Arbor, Transportation Research Institute.

[31] Jones, P. R., Ungewiss, J., Eichinger, P., Wörner, M., Crabb, D. P., & Schiefer, U. (2022). Contrast sensitivity and night driving in older people: Quantifying the relationship between visual acuity, contrast sensitivity, and hazard detection distance in a night-time driving simulator. Frontiers in human neuroscience, 16, 914459.

[32] Bullough, J. D., Fu, Z., & Van Derlofske, J. (2002). Discomfort and disability glare from halogen and HID headlamp systems (No. 2002-01-0010). SAE Technical Paper.

[33] Flannagan, M. J., Sivak, M., & Traube, E. C. (1994). Discomfort glare and brightness as functions of wavelength. University of Michigan, Ann Arbor, Transportation Research Institute.

[34] de Paiva, C. S. (2017). Effects of aging in dry eye. International ophthalmology clinics, 57(2), 47–64.

[35] Fujikado, T., Kuroda, T., Maeda, N., Ninomiya, S., Goto, H., Tano, Y., … & Mihashi, T. (2004). Light scattering and optical aberrations as objective parameters to predict visual deterioration in eyes with cataracts. Journal of Cataract & Refractive Surgery, 30(6), 1198–1208.

[36] Shandiz, J. H., Derakhshan, A., Daneshyar, A., Azimi, A., Moghaddam, H. O., Yekta, A. A., … & Esmaily, H. (2011). Effect of cataract type and severity on visual acuity and contrast sensitivity. Journal of ophthalmic & vision research, 6(1), 26.

[37] Zobor, D., Zobor, G., & Kohl, S. (2015). Achromatopsia: on the doorstep of a possible therapy. Ophthalmic research, 54(2), 103–108.

[38] Kohl, S., Jägle, H., Wissinger, B., & Zobor, D. (2018). Achromatopsia. In n GeneReviews®. University of Washington, Seattle, Seattle (WA). PMID: 20301591.

[39] Zeitz, C., Robson, A. G., & Audo, I. (2015). Congenital stationary night blindness: an analysis and update of genotype–phenotype correlations and pathogenic mechanisms. Progress in retinal and eye research, 45, 58–110.

[40] Reiss, B., & Ripperger, J. (2017). Laser technology in exterior lighting for vehicles. ATZ worldwide, 119(11), 54–59.

[41] Long, X., He, J., Zhou, J., Fang, L., Zhou, X., Ren, F., & Xu, T. (2015). A review on light-emitting diode based automotive headlamps. Renewable and Sustainable Energy Reviews, 41, 29–41.

[42] Reidenbach, H. D. (2009). Local susceptibility of the retina, formation and duration of afterimages in the case of Class 1 laser products, and disability glare arising from high-brightness light emitting diodes. Journal of laser applications, 21(1), 46–56.

[43] Veitch, J. A., & Miller, N. J. (2024). Effects of temporal light modulation on individuals sensitive to pattern glare. Leukos, 20(3), 310–346.

